# A Novel Multiplex qPCR Platform for Detection of *Mycobacterium tuberculosis* Drug Resistance Using Synthetic Mutation Standards and Fluorescence Quenching Probes

**DOI:** 10.1101/2025.06.10.25329335

**Authors:** Krishna H. Goyani, Daisy Patel, Shalin Vaniawala, Pratap N. Mukhopadhyaya

## Abstract

The emergence of multidrug-resistant (MDR) and extensively drug-resistant (XDR) *Mycobacterium tuberculosis* strains has intensified the need for rapid, accurate, and affordable molecular diagnostics. In this study, we report the development and analytical validation of a multiplex real-time PCR assay employing contact-quenching fluorescence detection, allele-specific primers, and 3′ blocked wild-type allele blockers for the detection of five clinically relevant resistance-associated mutations in the *rpoB, katG*, and *inhA* genes. Synthetic double-stranded linear DNA constructs bearing individual point mutations (*rpoB* codons 516, 526, 531; *katG* codon 315; and *inhA* (–)15 promoter) were engineered using the Splicing by Overlap Extension (SOEing) technique and confirmed by Sanger sequencing. These were used as positive controls in a background of wild-type genomic DNA from clinical isolates. The assay demonstrated high specificity and sensitivity, with detection limits ranging from 25 to 50 copies per reaction and effective mutant allele detection even in the presence of a 70-fold excess of wild-type DNA. The multiplex qPCR platform showed consistent amplification efficiency (92 to 104%), minimal cross-reactivity, and reliable performance even under PCR-inhibitory conditions. Inclusion of internal amplification controls and compatibility with multiple fluorescence channels support the robustness of the system. This study presents the first application of contact-quenching-based allele-specific PCR for MDR TB detection and offers a scalable, cost-effective alternative to commercial diagnostics. With its modular design and high analytical performance, the platform shows strong potential for deployment in decentralized settings for early drug resistance surveillance and patient management.

## Introduction

Tuberculosis (TB) continues to be a major global health issue in the 21st century, presenting a multifaceted challenge due to its enduring prevalence and the rise of drug-resistant variants. Although its incidence is gradually decreasing, TB still leads to around 8.6 million new infections and causes roughly 1.3 million deaths each year, with the majority of cases occurring among disadvantaged populations in low-income regions (Sulis et al., 2014). The World Health Organization classifies TB as a global public health emergency, noting that obstacles to effective disease management often stem more from socio-economic and political factors than from biological complexities (Ellner, 1997).

Current efforts to treat TB are hindered by the spread of multidrug-resistant (MDR) and extensively drug-resistant (XDR) strains of Mycobacterium tuberculosis (M. tb), which are resistant to existing treatment protocols. This challenge has driven the pursuit of novel drug therapies (Mabhula & Singh, 2019). Further complicating matters, co-infections involving multiple TB strains can obscure accurate diagnosis and hinder effective treatment, underscoring the urgent need for better diagnostic tools to improve clinical outcomes and control efforts (Cohen et al., 2012). Contemporary TB control frameworks promote individualized care, robust support infrastructures, and a commitment to research and innovation with the goal of eradicating TB within the coming decades (Sulis et al., 2014).

Enclosed environments, such as correctional facilities and areas with inadequate ventilation, play a critical role in facilitating TB transmission. This highlights the necessity for environmental interventions in such settings, especially since many heavily affected regions face shortages of healthcare professionals (Yates et al., 2016). The spread of M. tb within and beyond prison populations, particularly in crowded urban areas, calls for context-specific strategies aimed at breaking these transmission chains (Sanabria et al., 2023).

The rise of drug-resistant tuberculosis (TB), particularly in its multidrug-resistant (MDR-TB) and extensively drug-resistant (XDR-TB) forms, presents a formidable obstacle to effective disease management, making treatment both intricate and financially burdensome. MDR-TB exhibits resistance to key first-line drugs such as isoniazid and rifampin, whereas XDR-TB includes additional resistance to second-line medications, thereby complicating therapeutic approaches and diminishing treatment success (Iacobino et al., 2020; Jain & Dixit, 2008).

A major challenge in addressing drug-resistant TB lies in the prolonged and often toxic treatment regimens. These therapies are not only more demanding but are also associated with severe side effects, poor patient adherence, and limited cure rates. Compared to drug-sensitive TB, the treatment of MDR/XDR-TB requires extended durations, substantially higher costs, and more intricate drug protocols, often necessitating enhanced clinical support (Lange et al., 2014). This complexity is further exacerbated in patients living with HIV, resulting in poorer outcomes and increased strain on healthcare resources (Singh et al., 2020).

To mitigate these challenges, the World Health Organization (WHO) has advocated for an integrated approach focused on expanding rapid diagnostic capabilities, ensuring timely initiation of appropriate therapies, and reinforcing infection prevention strategies. These measures are intended to curb the transmission of resistant TB strains and address the socio-economic disparities that facilitate their spread (Matteelli et al., 2016). Nonetheless, the global burden of drug-resistant TB persists, primarily due to insufficient implementation of standardized guidelines and resource deficits, particularly in high-incidence regions (Migliori et al., 2012; Migliori et al., 2010).

Real-time PCR assays utilizing TaqMan probes have become essential tools for identifying drug resistance in *Mycobacterium tuberculosis*, the causative agent of tuberculosis (TB). These molecular diagnostics provide a fast and reliable means of detecting mutations linked to resistance against primary anti-TB medications.

Among these, the MeltPro TB/INH assay stands out as a closed-tube, dual-color real-time PCR test specifically developed to identify 30 known mutations linked to isoniazid resistance across several genetic loci in *M. tuberculosis*. Clinical evaluation of this assay demonstrated excellent performance, achieving 90.8% sensitivity and 96.4% specificity for the detection of isoniazid resistance. Furthermore, the assay supports rapid processing on a variety of real-time PCR platforms, offering efficiency and adaptability (Hu et al., 2014).

Another notable system is the Anyplex MTB/NTM MDR-TB assay, a multiplex PCR-based method that simultaneously detects the *Mycobacterium tuberculosis* complex and screens for multidrug-resistant strains. When tested on both pulmonary and extrapulmonary specimens, this assay exhibited perfect specificity (100%) in both contexts and achieved a sensitivity rate of 83.3% for isoniazid resistance detection in each sample category (Sali et al., 2015).

Contact quenching is a specific mechanism of fluorescence quenching that involves the direct interaction between a fluorophore and a quencher molecule, leading to a decrease in fluorescence intensity. Contact quenching requires molecular contact between the two entities for it to occur, which differentiates it from other quenching mechanisms like collisional (dynamic) quenching, where the quenching occurs due to transient collisions. In contact quenching, also known as static quenching, a non-fluorescent complex is formed between the fluorophore and the quencher. This is a result of the ground-state interaction, meaning the formation of this non-fluorescent complex occurs before the excitation of the fluorophore. In static quenching, the contact and resultant complex formation is integral to the quenching process, preventing the fluorophore from fluorescing (Quenching of Fluorescence, 2006).

In this study, we applied the principle of contact quenching to identify *Mycobacterium tuberculosis* mutations associated with drug resistance. To the best of our knowledge, this represents the first attempt to utilize this technology for detecting clinically significant mutations within the *M. tuberculosis* genome in a multiplexed format.

## Material and methods

### *Reference Strain and DNA* Extraction

*Mycobacterium tuberculosis* isolates lacking known resistance-associated mutations were procured from MicroCare Laboratory and TB Research Center (Surat, India). These wild-type isolates were confirmed to be mutation-free using Line Probe Assay (LPA) and GeneXpert MTB/RIF (Cepheid, USA) testing. Genomic DNA was extracted using the Wobble Base MTB DNA Extraction Kit (Wobble Base Bioresearch, Surat, India), following the manufacturer’s instructions. DNA quality and quantity were assessed using a LABMAN Digital Double Beam Spectrophotometer (U Tech Labs, Mumbai, India) and the samples were stored at −20°C until further analysis.

### Mutation Information and Synthetic Control Preparation

Clinically relevant resistance-associated mutations were selected based on standardized genome annotation and mutation databases, including the Tuberculosis Drug Resistance Mutation Database (TBDReaMDB) and WHO catalogue of mutations in *Mycobacterium tuberculosis* complex and their association with drug resistance (2021 version). These mutations spanned regions in key resistance-related genes such as *rpoB, katG*, and *inhA* promoter.

Synthetic linear double-stranded DNA fragments harbouring the desired mutations were generated using site-directed mutagenesis via the Splicing by Overlap Extension (SOEing) PCR method. These synthetic mutant templates were designed to mimic clinically observed single-nucleotide polymorphisms (SNPs) and used as positive controls. The integrity and identity of the introduced mutations were confirmed by Sanger sequencing of the final mutated double-stranded DNA constructs, ensuring sequence fidelity and accurate representation of clinically relevant variants.

### Design of Allele-Specific Primers and Fluorescent Probes

Custom allele-specific primers (ASPs) were designed such that the nucleotide position immediately before the 3′ terminus corresponded to the target SNP site. Each ASP was appended with a distinct 5′ tag sequence to enable differentiation during multiplex detection. Fluorescent universal probes (UPs), each bearing a unique fluorophore (e.g., FAM, HEX, Cy3, Cy5) at the 5′ end, were designed to hybridize specifically to these tags. A shared quencher oligonucleotide (Common-Q), modified with a 3′ Black Hole Quencher 1 (BHQ1), was used to suppress background signal via contact-mediated quenching of unbound probes. Primer and probe sets were computationally optimized to ensure minimal cross-reactivity, compatible melting temperatures (Tm), and favourable thermodynamic stability. Amplicon lengths were designed in the 100–200 bp range to facilitate efficient amplification and detection. Final primer combinations were evaluated for multiplex compatibility and potential dimer formation.

### Quantitative PCR and Cycling Conditions

Real-time PCR amplification was performed using either the QuantStudio 7 (Thermo Fisher Scientific, USA) equipped with multiple fluorescence detection channels compatible with the fluorophores used in this study. Each 15μL reaction mixture contained 1×PCR buffer with 3mM MgCl2, 0.2mM of each dNTP, 0.1μM of each allele-specific forward primer, 0.3μM reverse primer, 0.2μM of each universal fluorescent probe (UP), 0.5μM of the common quencher oligonucleotide (C-Q), 0.5μM of the wild-type allele blocker probe, 0.5 units of Taq DNA polymerase, and target DNA corresponding to ≥50 genomic copies or an equivalent quantity of synthetic template DNA.

Thermal cycling was conducted under the following conditions: 95°C for 2Omin; 10 pre-amplification cycles of 95°C for 10s, 55°C for 20s, and 68°C for 20s; followed by 30 amplification cycles of 95°C for 10s, 68°C for 30s, and 55°C for 30s, with fluorescence acquisition during the final step of each cycle.

### Design and Evaluation of Wild-Type Allele Blocker Probes

To enhance the specificity of real-time PCR detection of key multidrug-resistant (MDR) mutations in *Mycobacterium tuberculosis*, a series of wild-type allele-specific blocker probes were designed to target the most clinically relevant mutation hotspots. These included loci within the rpoB gene, specifically Asp435Val (GAC→GTC), His445Tyr (CAC→TAC) and Ser450Leu (TCG→TTG), which lie within the rifampicin resistance-determining region (RRDR). The katG gene was targeted at Ser315Thr (AGC→ACC), a well-characterized mutation conferring high-level isoniazid resistance. Additionally, the inhA promoter region was targeted at – 15C→T, a regulatory mutation that leads to increased expression of the InhA enzyme and contributes to isoniazid resistance.

These oligonucleotide blockers were engineered to anneal specifically to the wild-type sequence at each locus and were modified at the 3′end with a blocking moiety to prevent polymerase extension. This design effectively occupied the wild-type binding site and suppressed nonspecific amplification by limiting primer access to non-mutant templates.

Allele-specific primers corresponding to mutant variants were co-amplified in the presence of wild-type allele blocking probes using carefully optimized differential primer-to-blocker concentrations to enhance the selective detection of mutant alleles while minimizing amplification from wild-type templates. The wild allele blockers were chemically modified at their 3′ ends to render them extension-deficient, thereby preventing them from serving as primers in the PCR reaction. This ensured that they did not generate any amplification products in conjunction with the reverse primers present in the assay, while still effectively occupying the wild-type binding sites to suppress nonspecific wild allele mediated amplification. The blocker oligonucleotides were evaluated for their ability to inhibit wild-type signal and for compatibility with multiplex qPCR workflows. The specific target mutations addressed by each blocker probe are listed in Table 1. Importantly, the inclusion of the allele blockers did not inhibit the amplification of mutant templates and allowed for successful discrimination of the relevant resistance-associated mutations at rpoB codons 516, 526, and 531, katG codon 315, and inhA –15 (see Figure 1A and 1B for representative amplification profiles).

**Table 1:** List of allele-specific blocking primers designed to suppress wild-type amplification at key resistance-associated loci in the Mycobacterium tuberculosis genome. Each primer selectively binds to the wild-type sequence and is 3′-modified to prevent extension during PCR, enabling specific amplification of mutant alleles. The table includes the oligonucleotide name, functional description, primer orientation, and the specific wild-type mutation site targeted for blocking.

| Name of the oligonucleotide primer | Description | Type of primer | Mutation blocked by the by the primer |
| --- | --- | --- | --- |
| KHG17072020-A | Allele blocker for rpoB gene mutation | Forward primer | GAC (GAC→GTC) rpoB gene; codon # 516 |
| KHG17072020-B | Allele blocker for rpoB gene mutation | Forward primer | **CAC** (CAC→TAC) rpoB gene; codon # 526 |
| KHG17072020-C | Allele blocker for rpoB gene mutation | Forward primer | TCG (TCG→TTG) rpoB gene; codon # 531 |
| HG17072020-N | Allele blocker for katG gene mutation | Forward primer | AGC (AGC→ACC) katG gene; codon # 315 |
| KHG17072020-U | Allele blocker for inhA gene mutation | Forward primer | inhA-15 position [C] in C→T |

**Figure 1:**
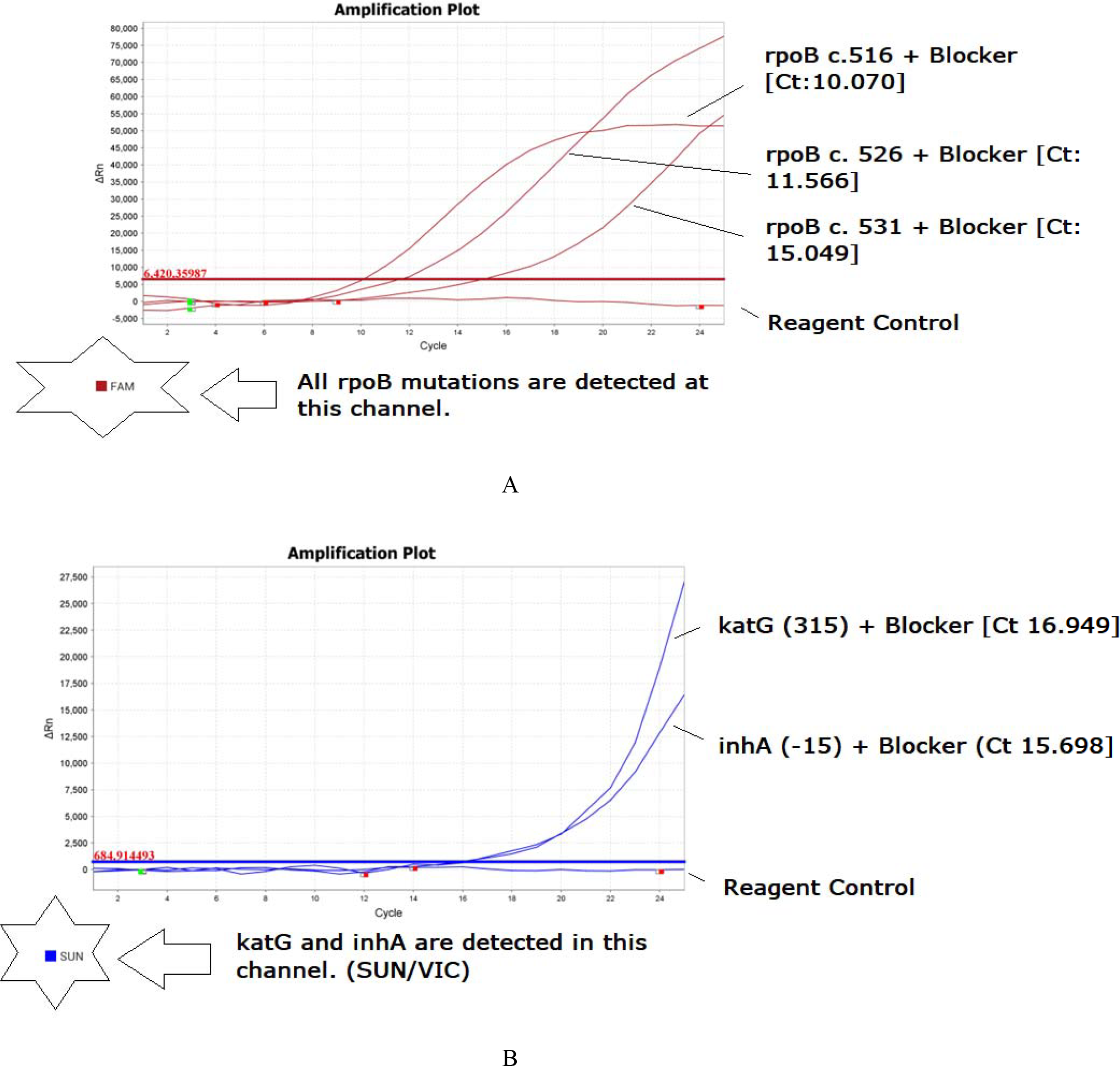
Amplification plots demonstrating successful detection of drug resistance-associated mutations in *Mycobacterium tuberculosis* using allele-specific real-time PCR. Panel A shows amplification of rpoB gene mutation targets using FAM-labelled probes, while Panel B shows amplification of katG and inhA mutation targets using SUN-labelled probes. Each reaction incorporated allele-specific primers, 3′-blocked wild-type allele blockers, fluorophore-labelled probes, and a common quencher probe employing a contact-quenching mechanism for sign discrimination.

All oligonucleotide primers, wild-type allele blocking probes, and dye/quencher-labeled contact-quenching probes used in this study were custom-designed using a proprietary algorithm developed at Wobble Base Bioresearch (Surat, India). The designs were specifically optimized for the selective detection of clinically significant mutations in the *Mycobacterium tuberculosis* genome. Each probe was engineered to operate effectively in a multiplex format utilizing a contact-quenching mechanism, thereby enhancing allele-specific signal discrimination. Due to intellectual property considerations, the exact sequences of the primers, allele blockers, and probes are not disclosed in this manuscript. However, all components were rigorously validated for analytical performance, including specificity, sensitivity, and compatibility with real-time PCR, and are available for academic collaboration under an appropriate material transfer agreement (MTA).

### Ethics Statement

This study utilized de-identified DNA extracted from a single sputum sample, which was collected by Microcare Laboratory and TB Research Center, Surat, under standard diagnostic procedures. The sample was used exclusively for non-clinical laboratory applications, specifically for site-directed mutagenesis and in vitro assay development. The study protocol was reviewed and approved by the Wobble Base Bioresearch Ethics Committee (Approval No. WBBPL/EC/J2021/001), in accordance with applicable ethical standards and the Declaration of Helsinki. No identifiable patient information was accessed or recorded, and no direct human participation was involved.

## Results

### Synthesis and Sequence Validation of Synthetic Mutation Controls

To ensure reliable detection and discrimination of multidrug-resistant (MDR) mutations in *Mycobacterium tuberculosis*, synthetic DNA controls were engineered to contain specific resistance-associated mutations within the *rpoB, katG*, and *inhA* genes. These included codon substitutions at *rpoB* positions c.516 (GAC→GTC), c.526 (CAC→TAC), and c.531 (TCG→TTG); *katG* codon 315 (AGC→ACC); and the *inhA* promoter region at –15 (C→T). Synthetic control generation was accomplished using the Splicing by Overlap Extension (SOEing) method, a restriction site-independent, PCR-based technique for in vitro mutagenesis and DNA fragment recombination.

For each target mutation, two overlapping sub-amplicons were amplified using outer primers (non-mutagenic) in combination with internal primers carrying the desired base substitution. The resulting overlapping fragments were denatured, annealed, and extended in a subsequent PCR reaction using the outer primer pair to generate a full-length linear product harbouring the specific mutation. The list of primers and respective amplicon sizes for all mutation targets is provided in Table 2.

**Table 2:**
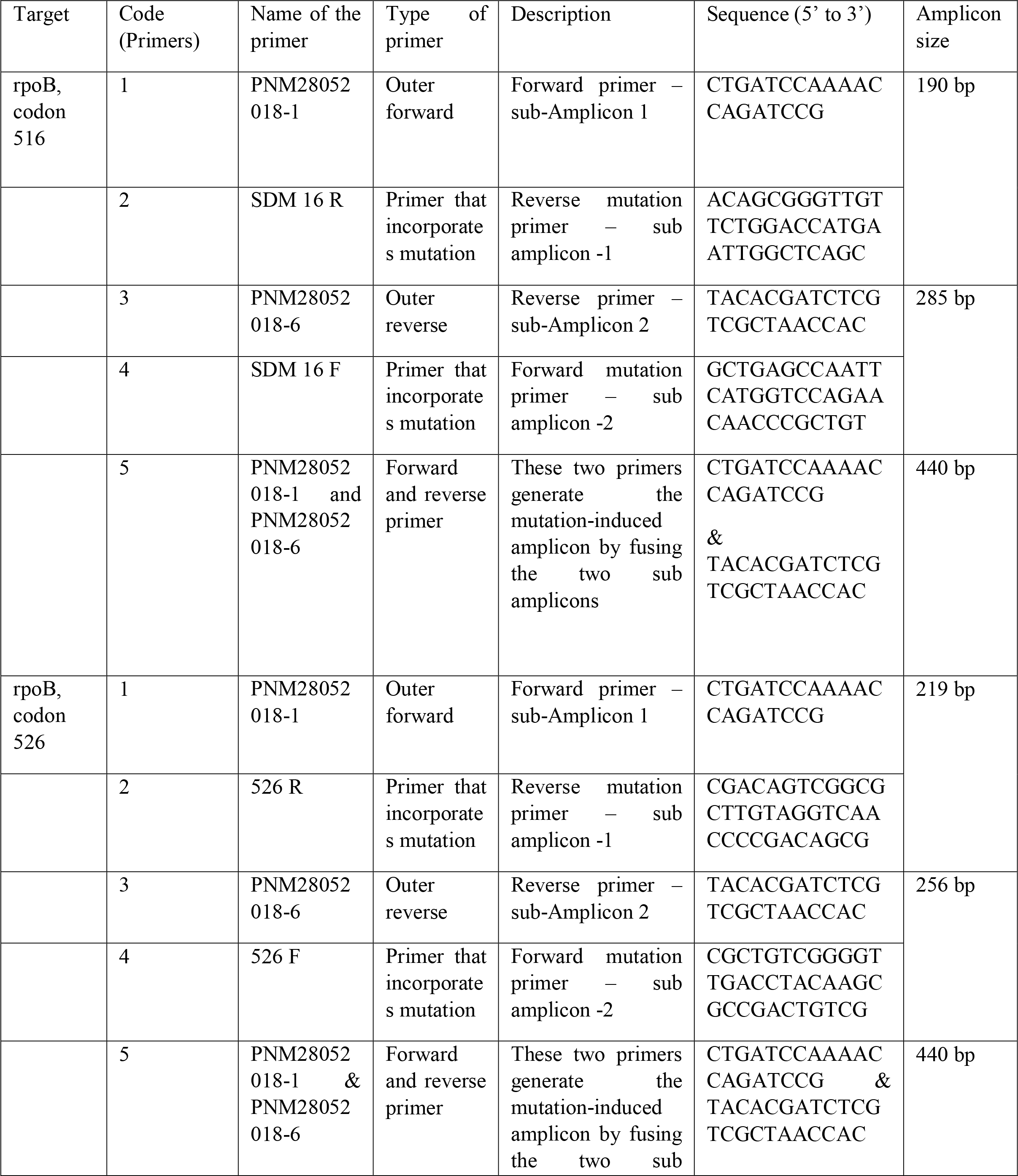
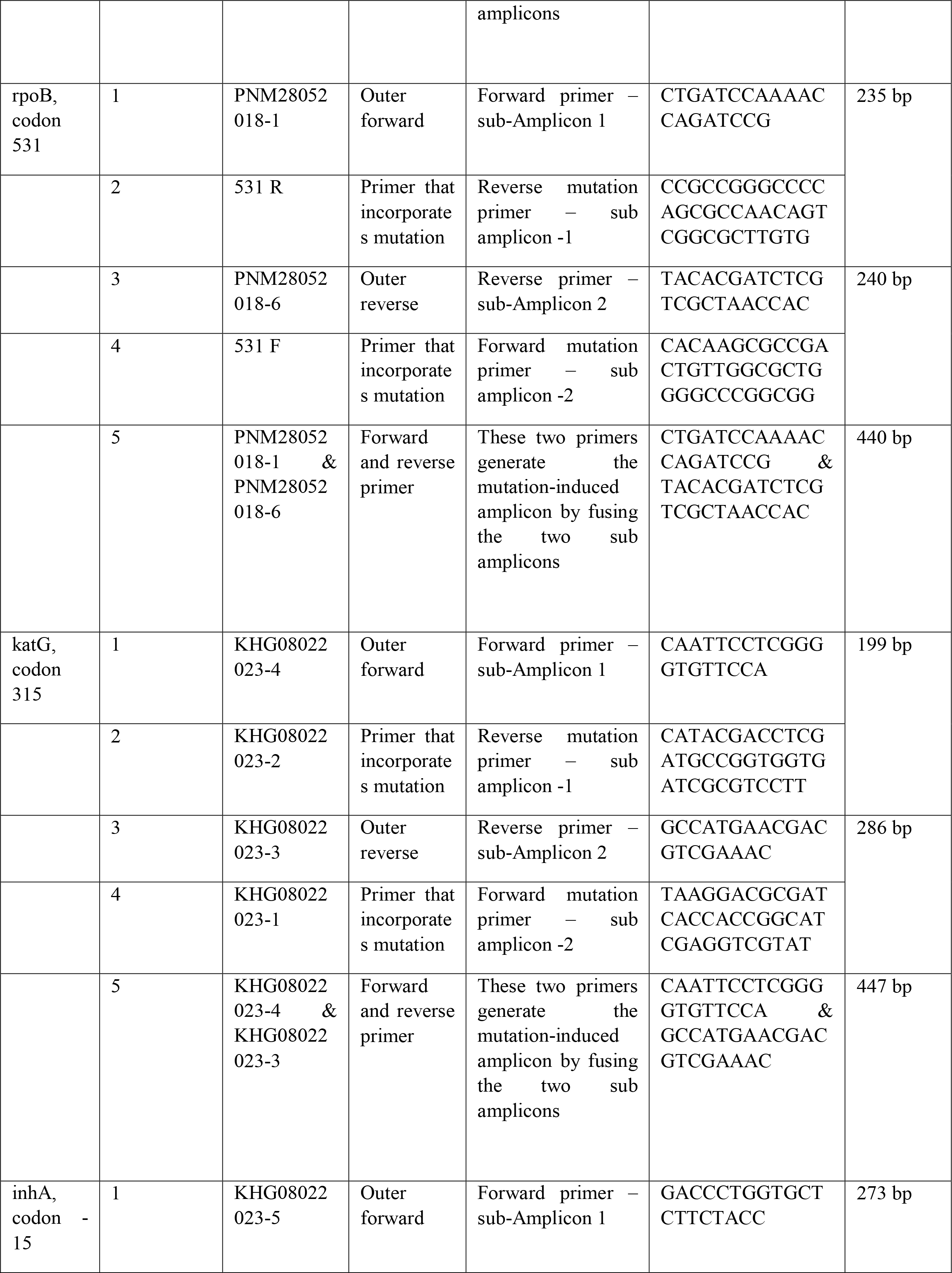
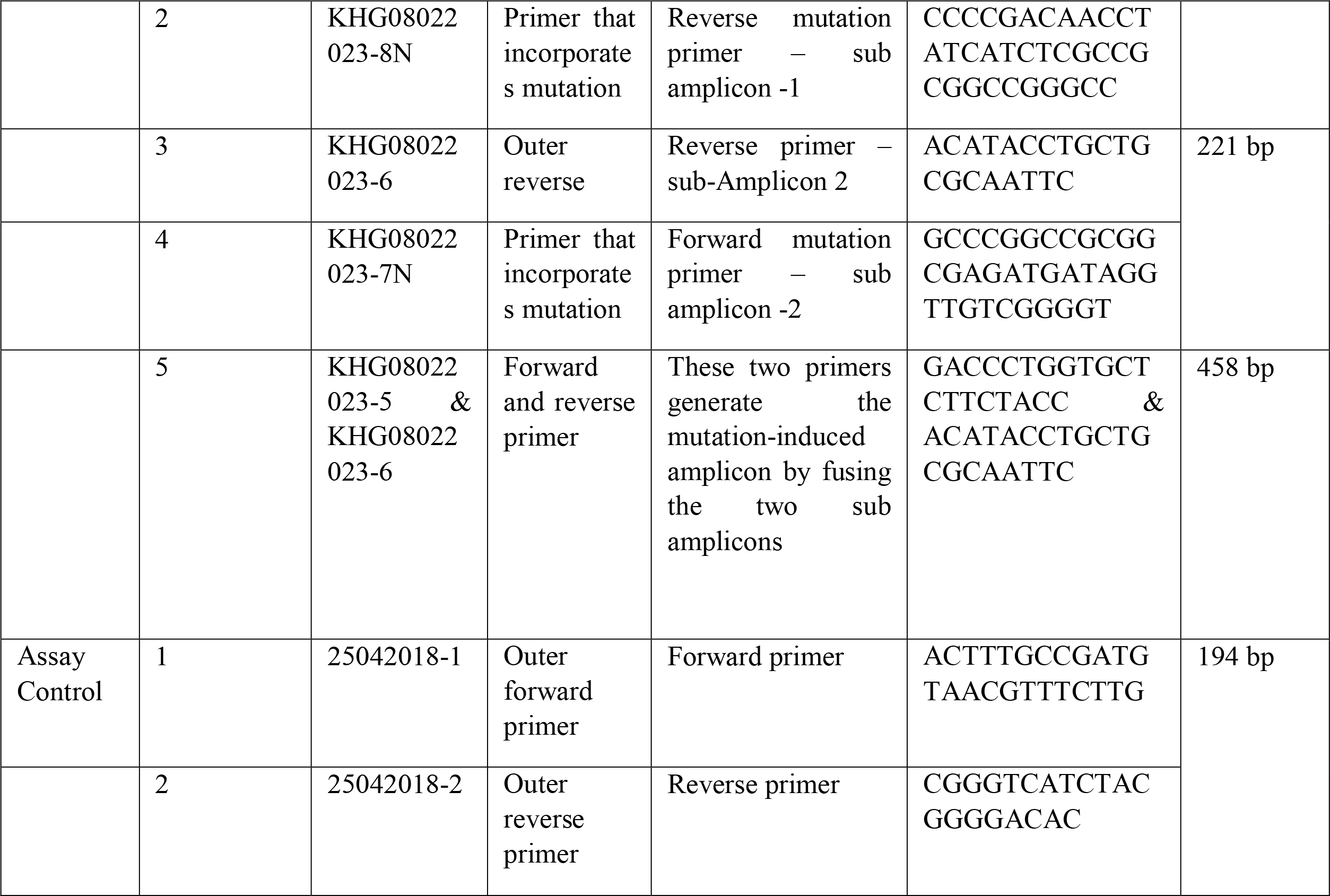
List of primers used for the synthesis of mutant DNA constructs targeting key drug resistance-associated loci in *Mycobacterium tuberculosis*. For each target mutation (rpoB codons 516, 526, 531; katG codon 315; and inhA –15 promoter), two overlapping sub-amplicons were generated using outer primers and site-directed mutagenesis (SDM) primers carrying the desired single-nucleotide polymorphisms (SNPs). The sub-amplicons were fused in a secondary PCR to produce the final mutation-containing linear DNA constructs. The table details the primer codes, types, descriptions, sequences (5′–3′), and resulting amplicon sizes. An additional control primer set was included for internal assay validation.

Electrophoretic analysis confirmed successful amplification of sub-amplicons and full-length products. Figure 2 displays the sub-amplicons for *rpoB* mutations at codons 516, 526, and 531, while Figure 3 presents the final fused amplicons representing the complete mutant constructs. The integrity and mutation incorporation within the *rpoB* synthetic controls were verified via capillary sequencing, with the electropherograms confirming accurate introduction of the GAC→GTC (c.516), CAC→TAC (c.526), and TCG→TTG (c.531) substitutions (Figures 4,5 and 6, respectively). Similarly, the two sub-amplicons for *katG* codon 315 and *inhA* –15 promoter mutation were successfully generated (Figure 7, panels A and B), followed by fusion into a single synthetic fragment (Figure 7, panel C). Capillary sequencing validated the presence of the AGC→ACC mutation in *katG* (Figure 8) and the –15C→T mutation in the *inhA* promoter region (Figure 9).

**Figure 2.**
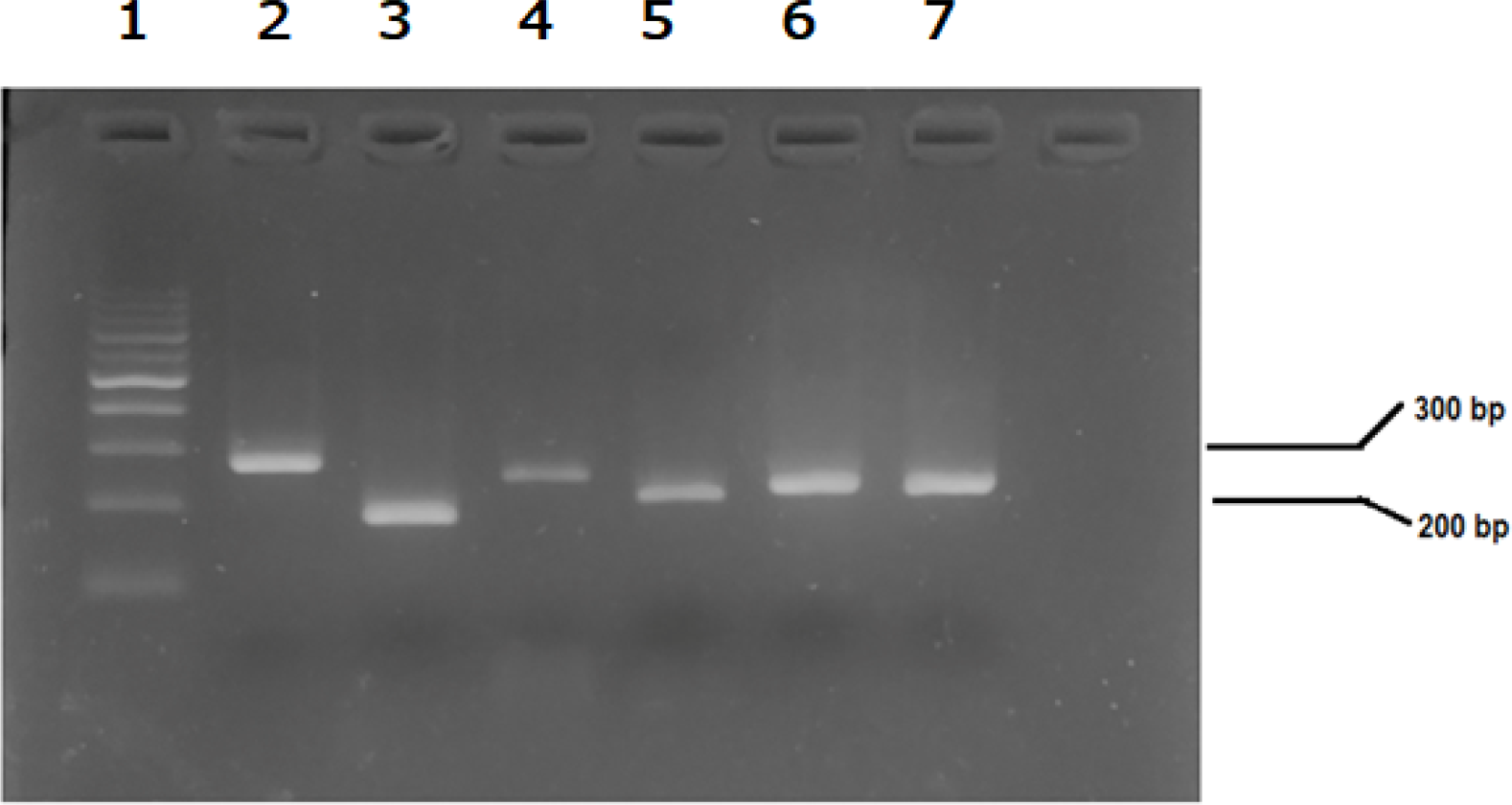
Agarose gel electrophoresis showing sub-amplicons generated for the synthesis of rpoB mutation controls used in this study. Lane 1: DNA ladder; Lanes 2 and 3: sub-amplicons corresponding to the rpoB codon 51 (GAC→GTC) mutation; Lanes 4 and 5: sub-amplicons for rpoB codon 526 (CAC→TAC); Lanes 6 and 7: sub-amplicons for rpoB codon 531 (TCG→TTG). The expected sizes of these PCR fragments are detailed in Table (last column), with the top two rows listed for each AMR target.

**Figure 3.**
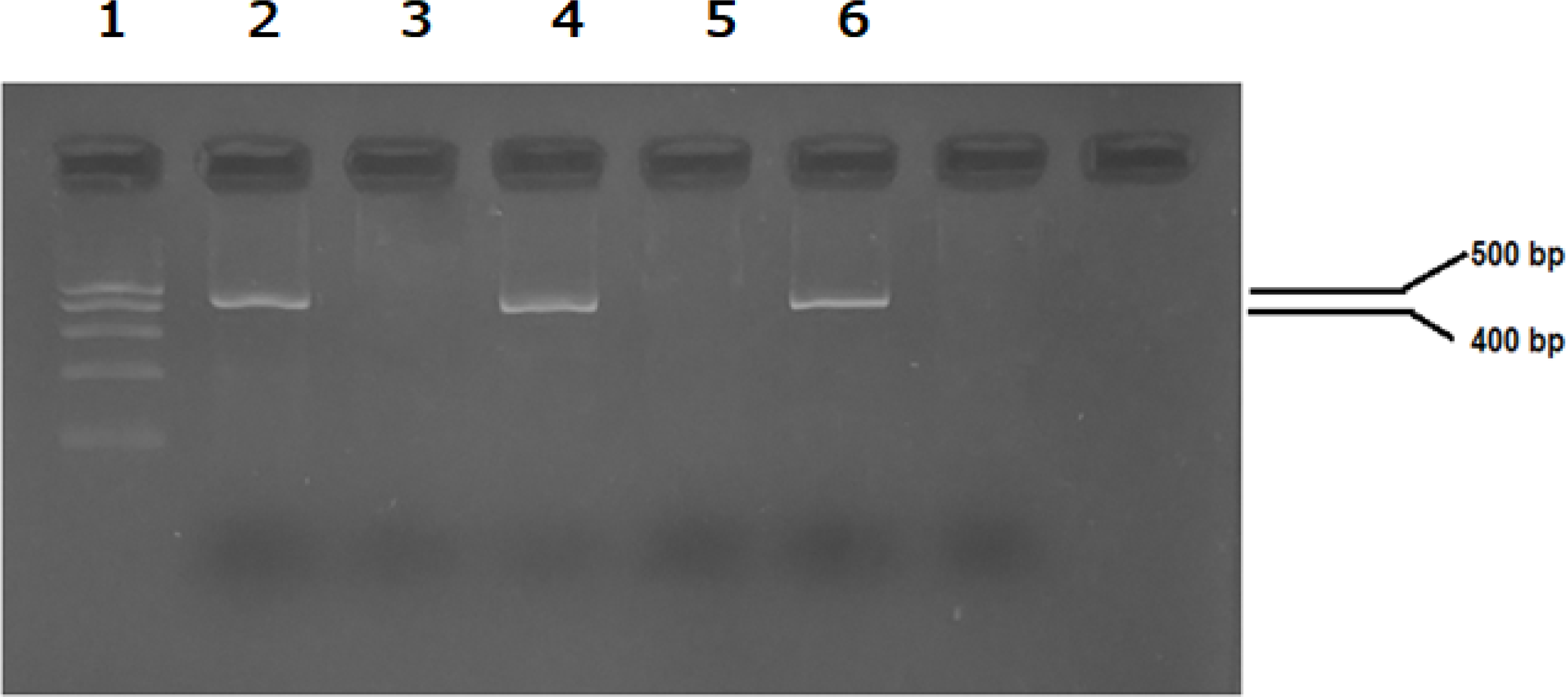
Generation of mutation-induced full-length single-stranded DNA constructs using the Splicing by Overlap Extension (SOEing) PCR technique. Lane 1: DNA ladder; Lane 2: final amplicon for rpoB codon 516 (GAC→GTC); Lane 4: final amplicon for rpoB codon 526 (CAC→TAC); Lane 6: final amplicon for rpoB codo 531 (TCG→TTG). The expected sizes of these mutation-containing PCR products are listed in Table 2 (last column), specifically in the third row from the top for each corresponding AMR target.

**Figure 4.**
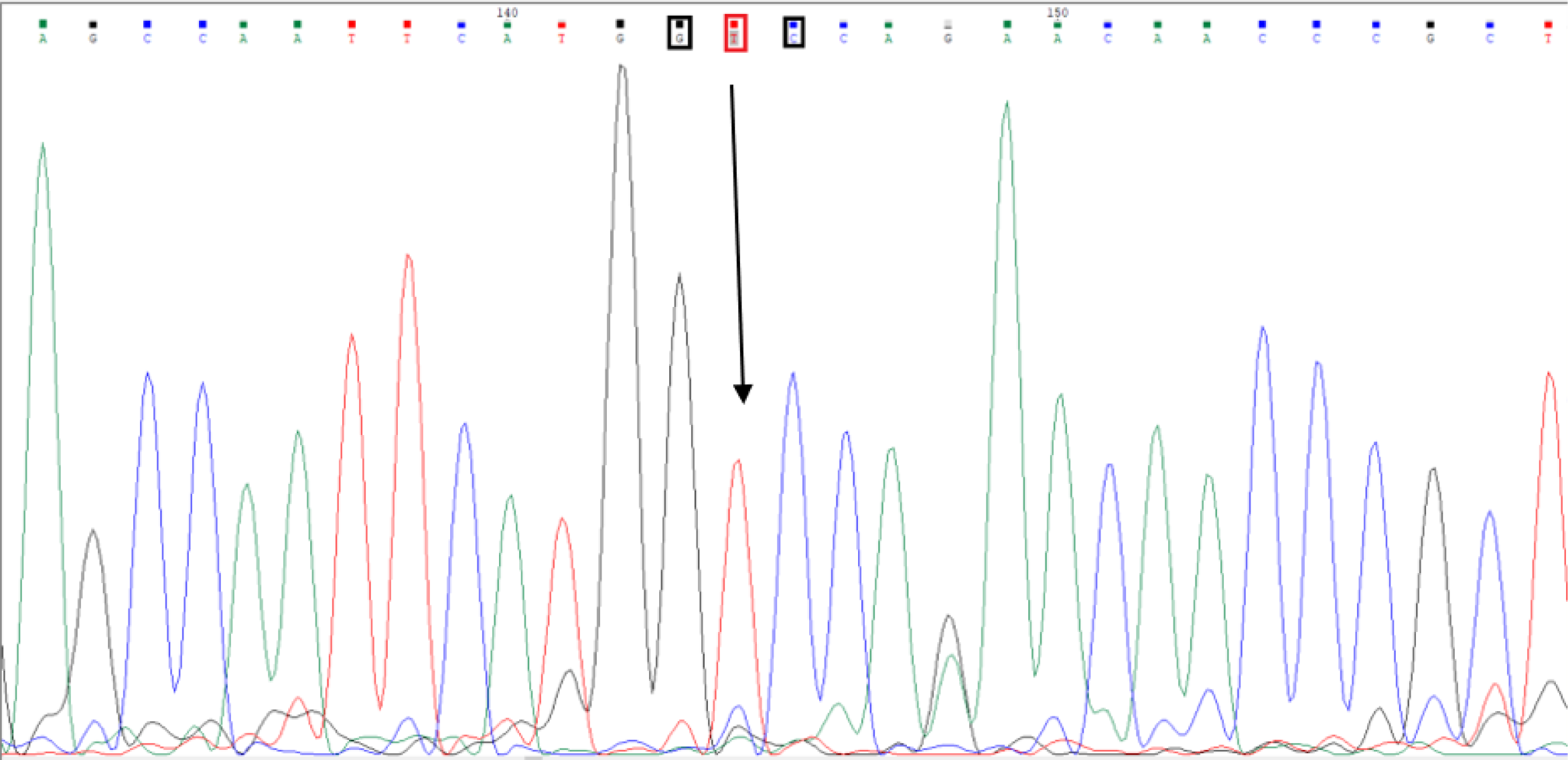
Representative electropherogram confirming successful incorporation of the rpoB codon 516 mutation (GAC→GTC) within the SOEing-derived synthetic control DNA. The mutation site is clearly visible, validating th accuracy of the Splicing by Overlap Extension (SOEing) PCR-based mutagenesis and the fidelity of the final synthetic construct.

**Figure 5.**
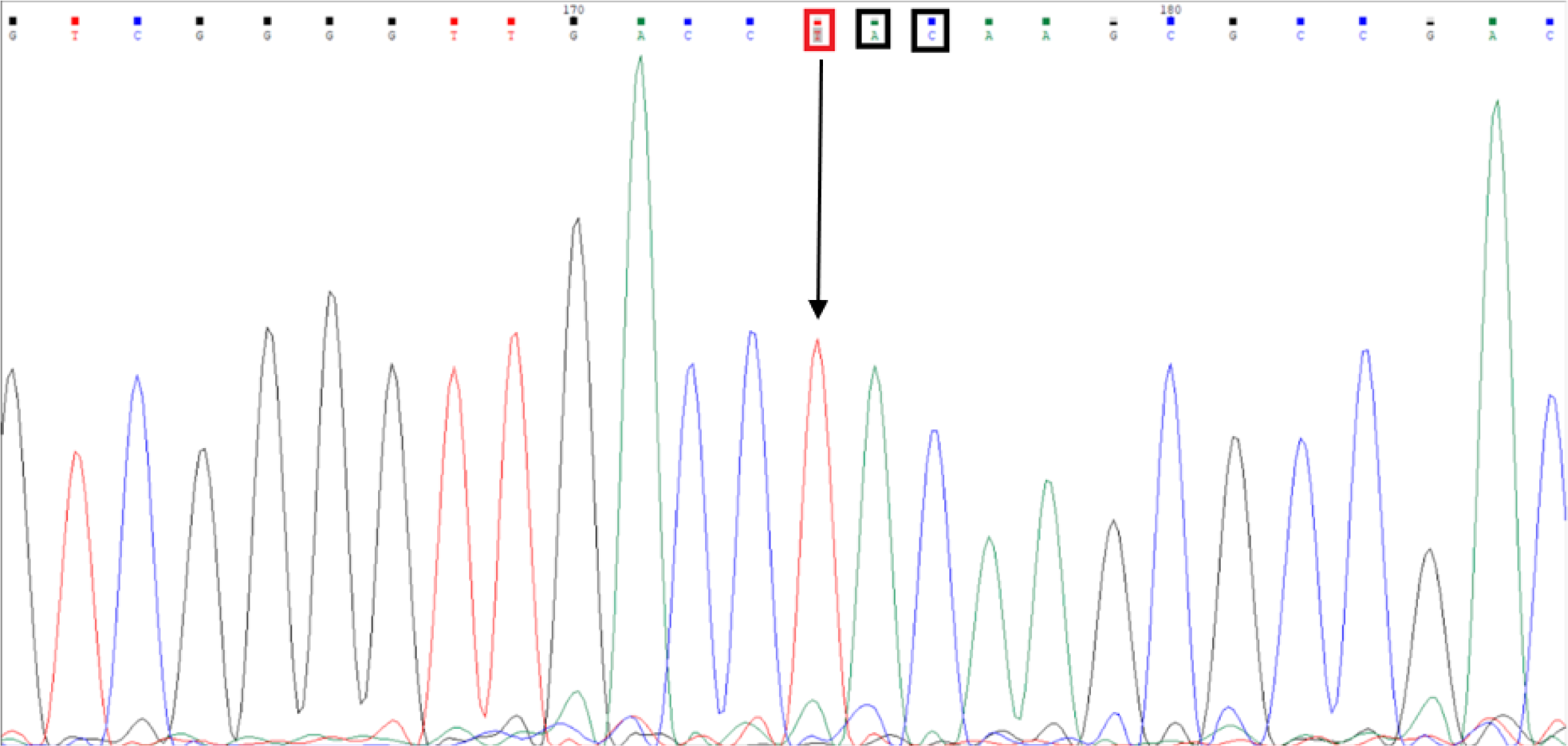
Electropherogram confirming successful incorporation of the rpoB codon 526 mutation (CAC→TAC) within the SOEing-derived synthetic control DNA. The sequence trace clearly shows the intended base substitution, validating precise mutagenesis and high-fidelity synthesis of the mutant construct.

**Figure 6.**
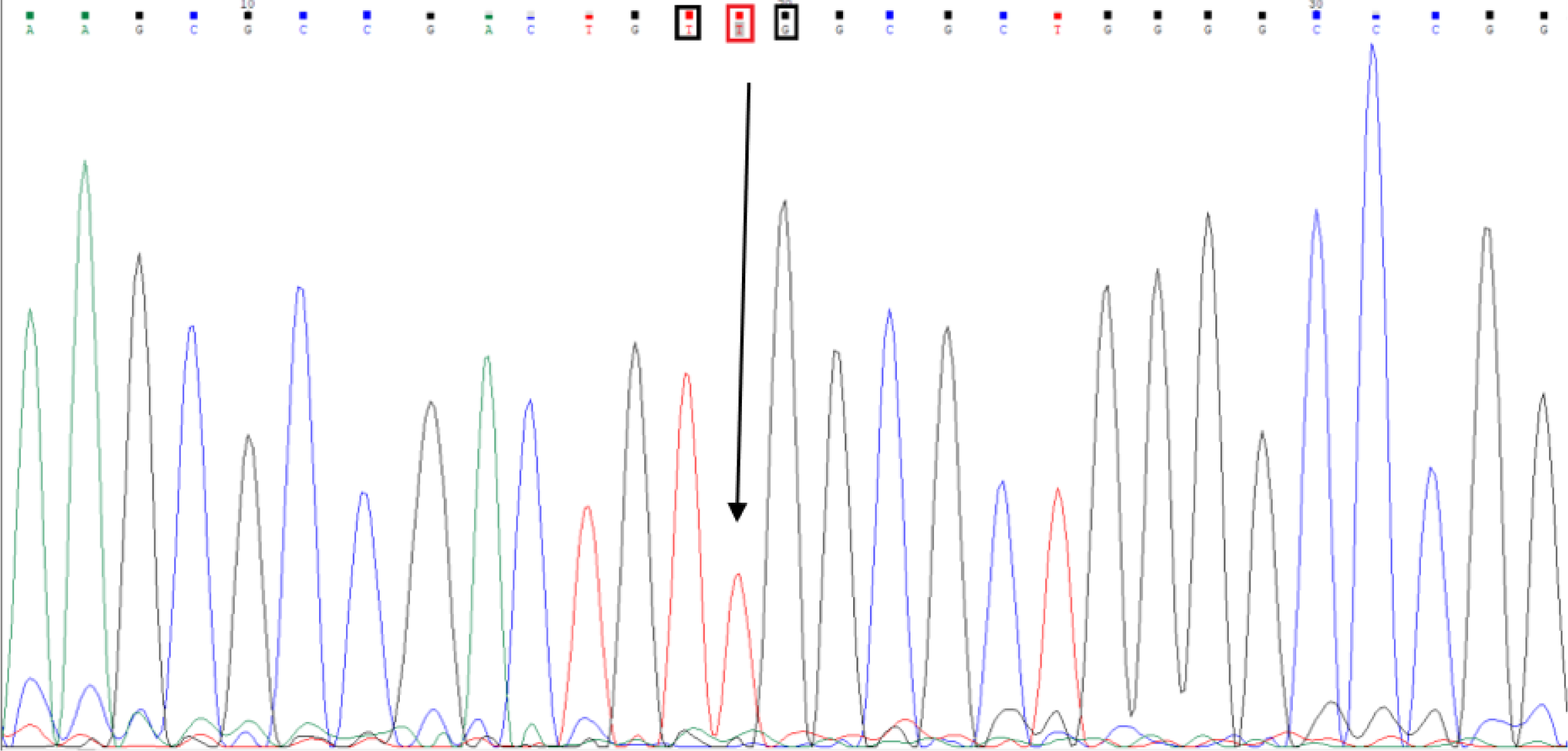
Electropherogram confirming successful incorporation of the rpoB codon 531 mutation (TCG→TTG) within the SOEing-derived synthetic control DNA. The highlighted base substitution demonstrates accurate site-directed mutagenesis and validates the fidelity of the final synthetic construct.

**Figure 7.**
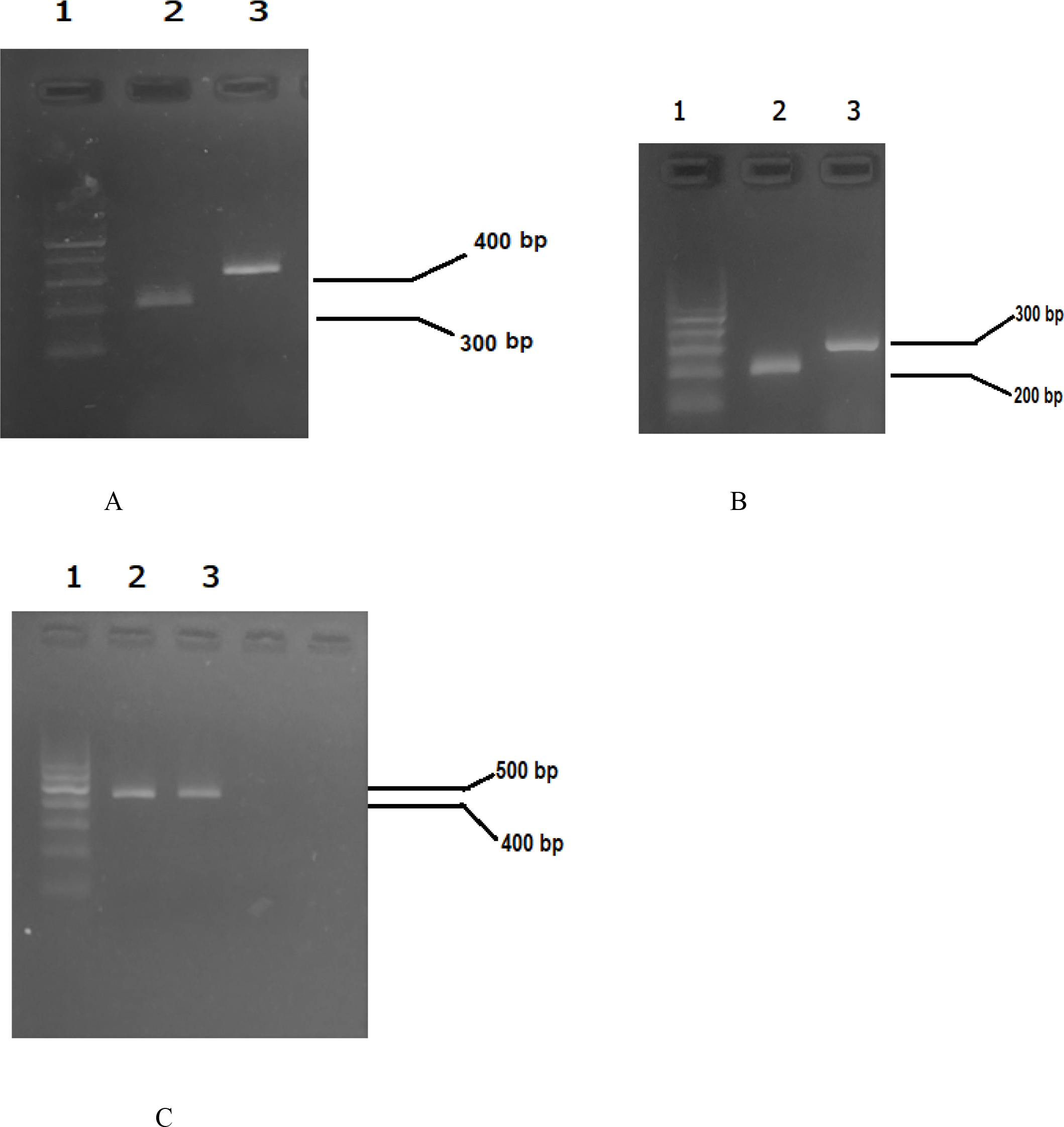
Agarose gel electrophoresis showing sub-amplicons and final reconstructed synthetic controls for katG (codon 315) and inhA (–15 promoter) mutation targets. Panel A: Lane 1: DNA ladder; Lanes 2 and 3: sub-amplicons generated for the katG codon 315 (AGC→ACC) mutation; Panel B: Lane 1: DNA ladder; Lanes 2 and 3: sub-amplicons for the inhA –15 (C→T) promoter mutation; Panel C: Lane 1: DNA ladder; Lanes 2 and 3: full-length reconstructed linear DNA fragments harbouring the respective mutations for katG and inhA, assembled using SOEing PCR. The expected amplicon sizes are provided in Table 2 (last column, third row for each AMR target).

**Figure 8.**
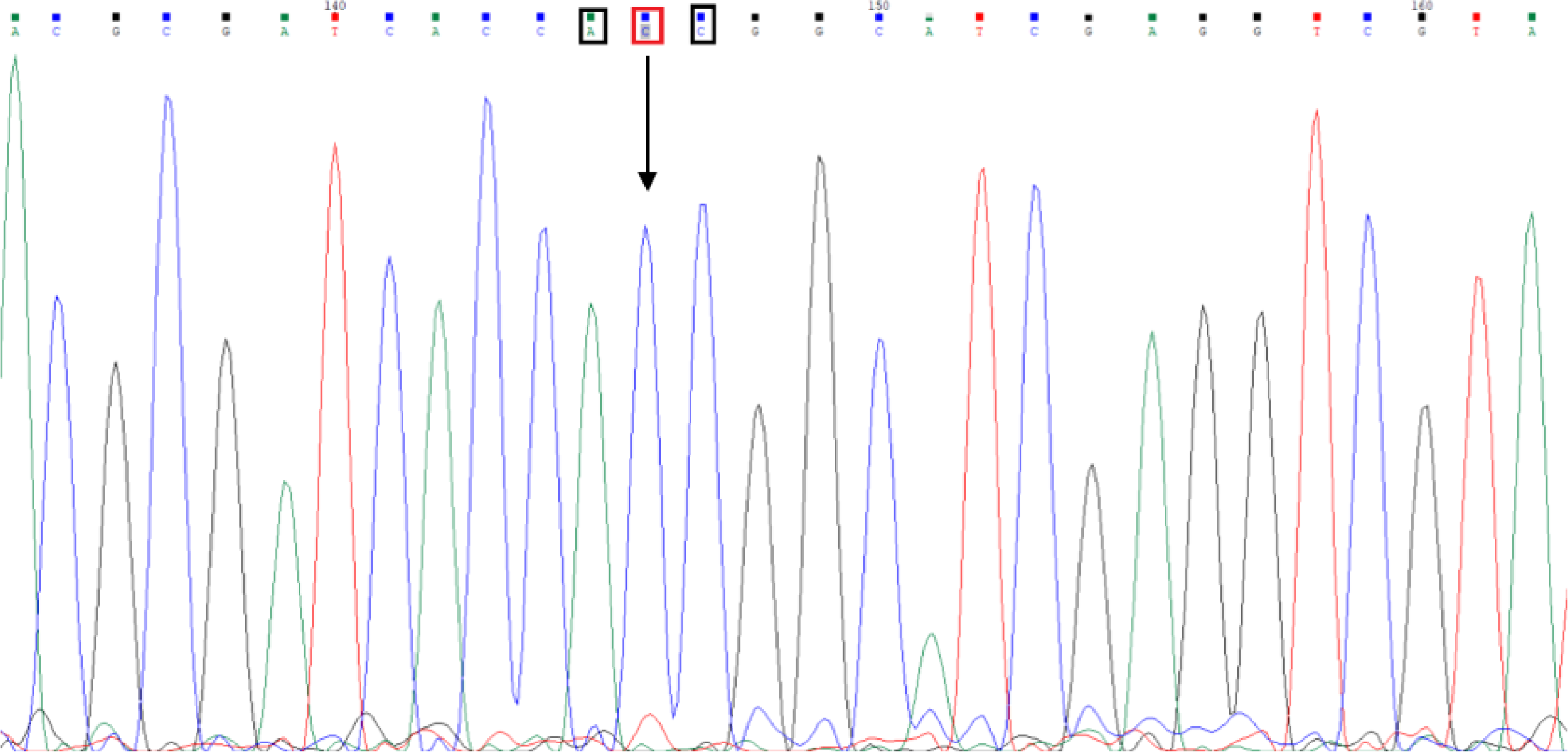
Electropherogram confirming successful incorporation of the katG codon 315 mutation (AGC→ACC) within the SOEing-derived synthetic control DNA. The sequence trace clearly shows the intended base substitution, validating precise site-directed mutagenesis and the fidelity of the synthetic construct.

**Figure 9.**
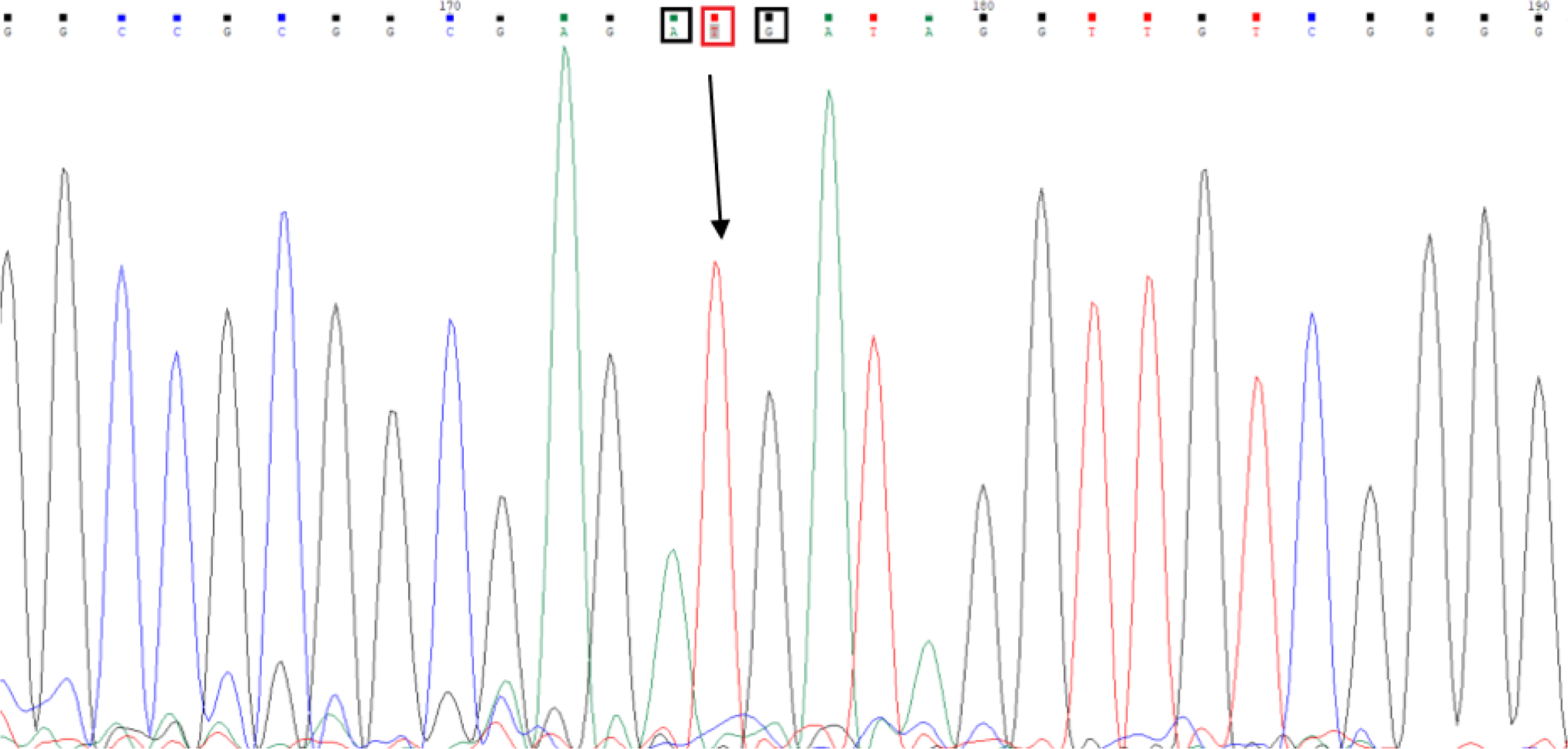
Electropherogram confirming successful incorporation of the inhA promoter –15 mutation (C→T) within the SOEing-derived synthetic control DNA. The highlighted base substitution verifies the accuracy of site-directed mutagenesis and the sequence integrity of the final synthetic construct.

These synthetic constructs serve as definitive mutation-specific templates and were used as controls for downstream real-time PCR assay development and validation.

### Validation of Wild-Type Allele Blocker Probes in Real-Time PCR Assay

To assess the functional performance of the wild-type allele-specific blocker probes designed for selective amplification of mutant alleles, real-time PCR assays were performed targeting the clinically significant mutations within the *rpoB* (codons 516, 526, and 531), *katG* (codon 315), and *inhA* (–15 promoter region) genes. Each assay included mutant allele-specific primers, dual-labeled fluorescent detection probes, a common quencher oligonucleotide, and the respective wild-type blocker probes rendered extension-deficient by 3′ modification.

The results demonstrated that the inclusion of allele blockers did not inhibit PCR efficiency. Robust amplification was observed across all mutation targets, confirming that the blocker probes effectively suppressed non-specific amplification from wild-type templates while permitting specific detection of mutant alleles. Amplification of *rpoB* mutation targets is shown in Figure 1A, where FAM-labeled probes were used for real-time detection. Similarly, Figure 1B illustrates successful amplification of *katG* and *inhA* mutations using SUN-labeled probes under multiplex conditions.

These findings validate the functional compatibility of the blocker probes with the contact-quenching detection system and support their role in enhancing the specificity of allele-specific real-time PCR for drug resistance detection in *M. tuberculosis*.

### Assay Validation Using Synthetic Controls

To evaluate the analytical performance and specificity of the developed real-time PCR assays, synthetic DNA templates representing each of the five clinically relevant multidrug-resistant (MDR) *Mycobacterium tuberculosis* (MTB) mutations were used as positive controls. These included mutations in the *rpoB* gene at codons 516 (GAC→GTC), 526 (CAC→TAC), and 531 (TCG→TTG); *katG* at codon 315 (AGC→ACC); and the *inhA* promoter region at position –15 (C→T). In each reaction, wild-type *M. tuberculosis* genomic DNA (confirmed to be mutation-free at these loci) was mixed in a ratio of approximately 70:1 (wild-type: mutant) with the respective synthetic control, simulating low-abundance mutant alleles in a clinical background.

An internal amplification control (IAC) derived from a non-MTB target sequence was included in each reaction and consistently detected in the Cy5 channel, validating PCR efficiency and excluding inhibition. Each synthetic control assay was tested independently in parallel with a no-template control (NTC) to assess specificity and rule out reagent contamination.

The real-time PCR profiles are shown in Figure 10, with amplification traces generated on the QuantStudio 5 platform (Panels A1–G1). Parallel testing was conducted using the GeneXpert MTB/RIF system (Panels A2–E2) to confirm the wild-type status of the background genomic DNA employed in the synthetic control reactions. Figure 10A1 illustrates amplification of the *rpoB* c.516 mutant control alongside the internal amplification control (IAC), with GeneXpert confirming wild-type status at this locus (Figure 10A2). In Figure 10B1, detection of the *rpoB* c.526 mutant control is shown, and wild-type status was similarly validated by GeneXpert (Figure 10B2). Successful amplification of the *rpoB* c.531 mutant control is depicted in Figure10C1, with wild-type confirmation by GeneXpert (Figure10C2). Figure10D1 presents detection of the *katG* Ser315Thr mutant control, with GeneXpert verification of the wild-type background (Figure10D2). Amplification of the *inhA* –15 mutant synthetic template is shown in Figure10E1, and wild-type status was confirmed by GeneXpert (Figure10E2). Figure10F demonstrates the multiplex detection of both *rpoB* c.516 and *katG* 315 mutations within a dual-mutant synthetic control. Figure10G highlights assay performance in the presence and absence of PCR inhibitors, specifically EDTA at a final concentration of 75mM, thereby demonstrating the robustness of the system under inhibitory conditions. The template details used in each reaction are summarized in Table3. Collectively, these results confirm that the assays accurately and sensitively detect each mutation target, maintain specificity even in multiplexed formats, and remain functional in the presence of excess wild-type DNA or inhibitory substances.

**Figure 10.**
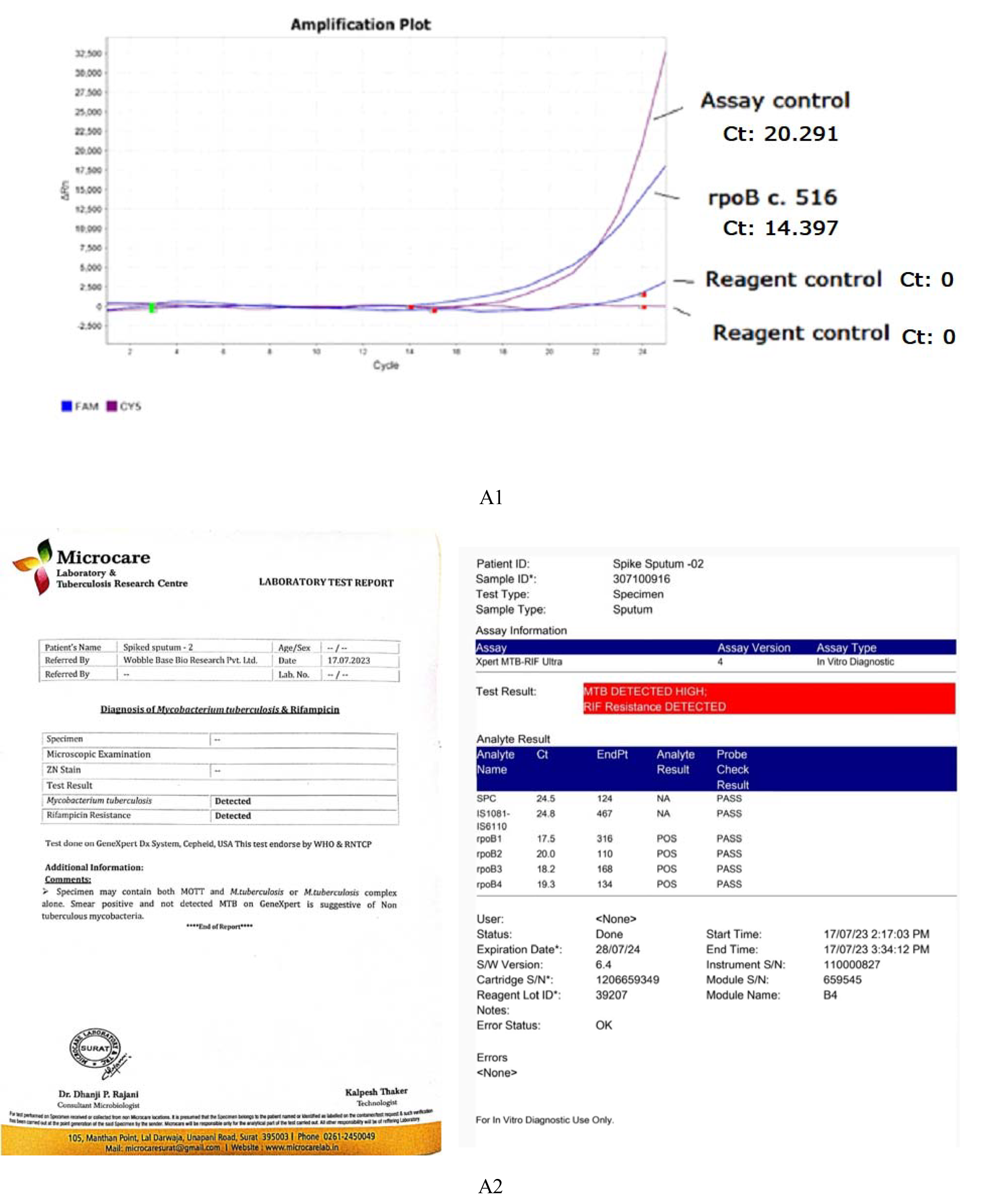
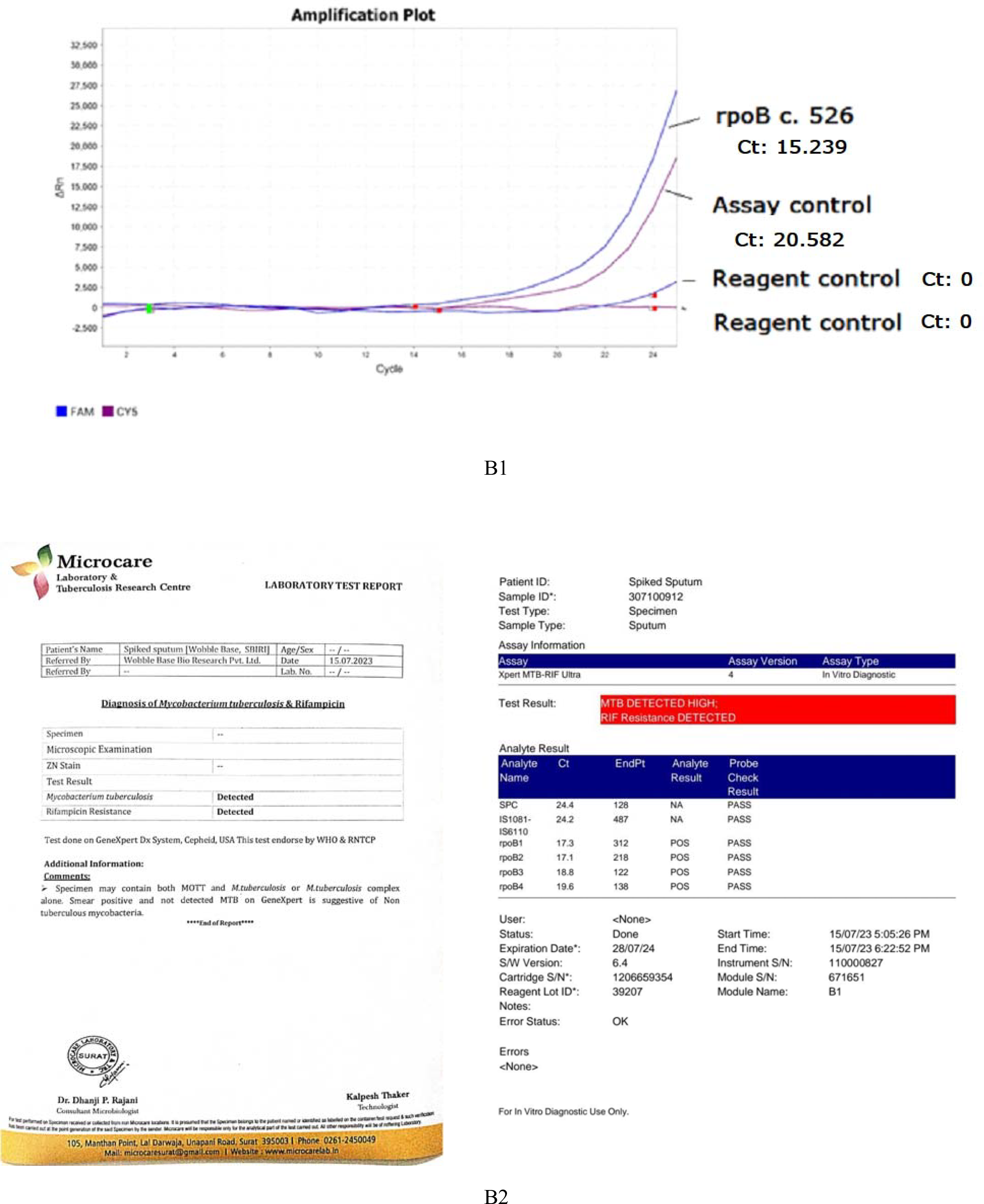
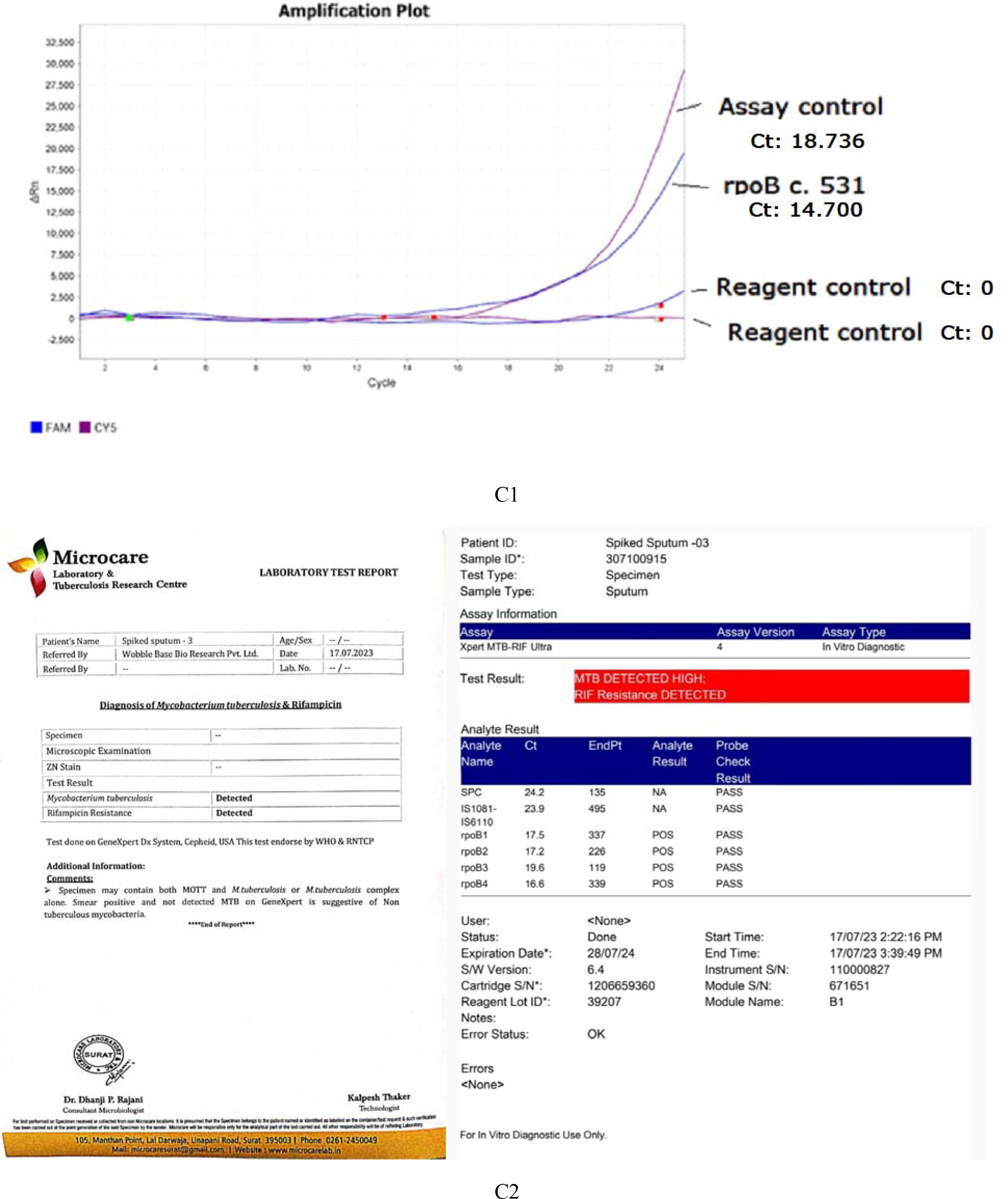
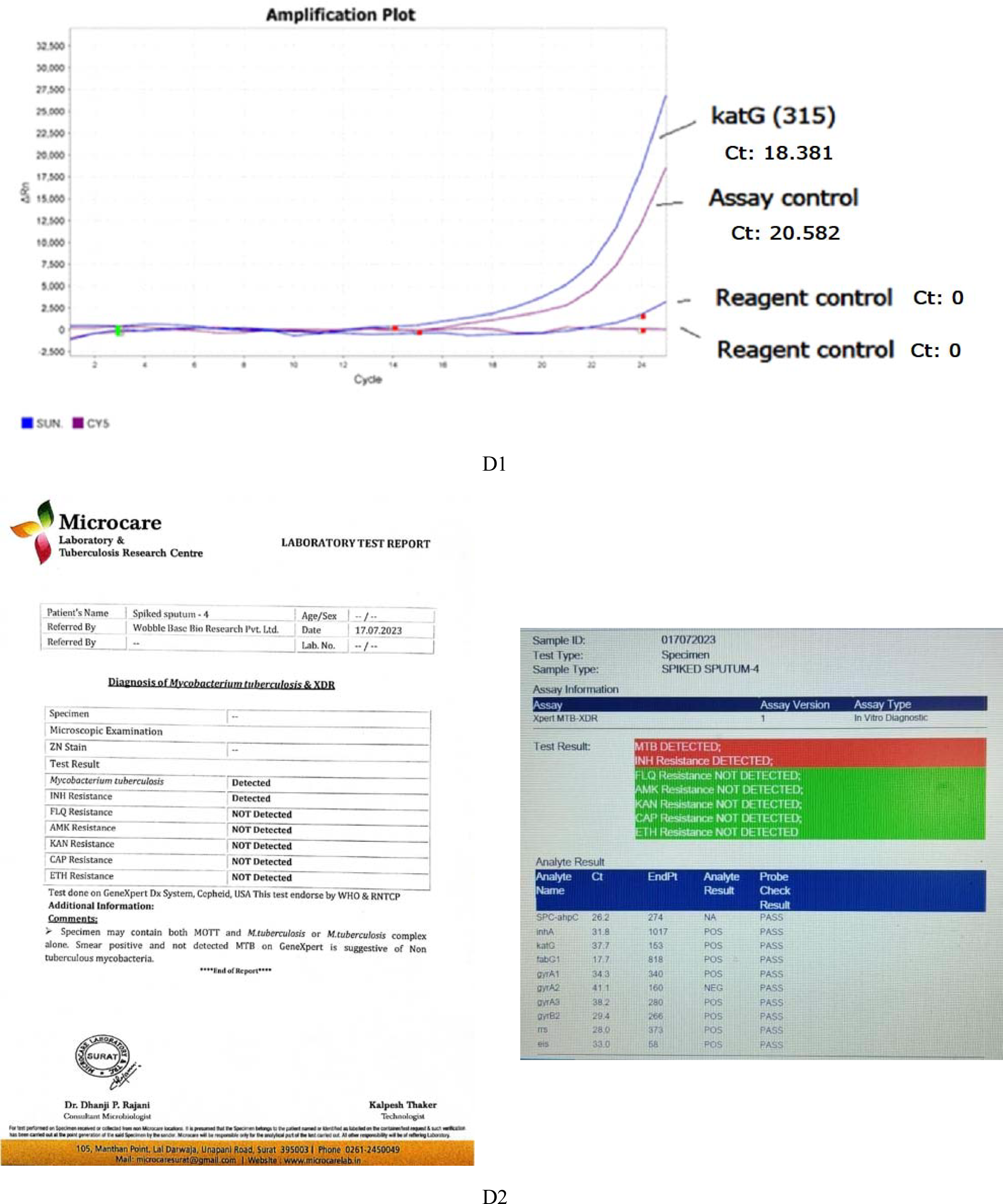
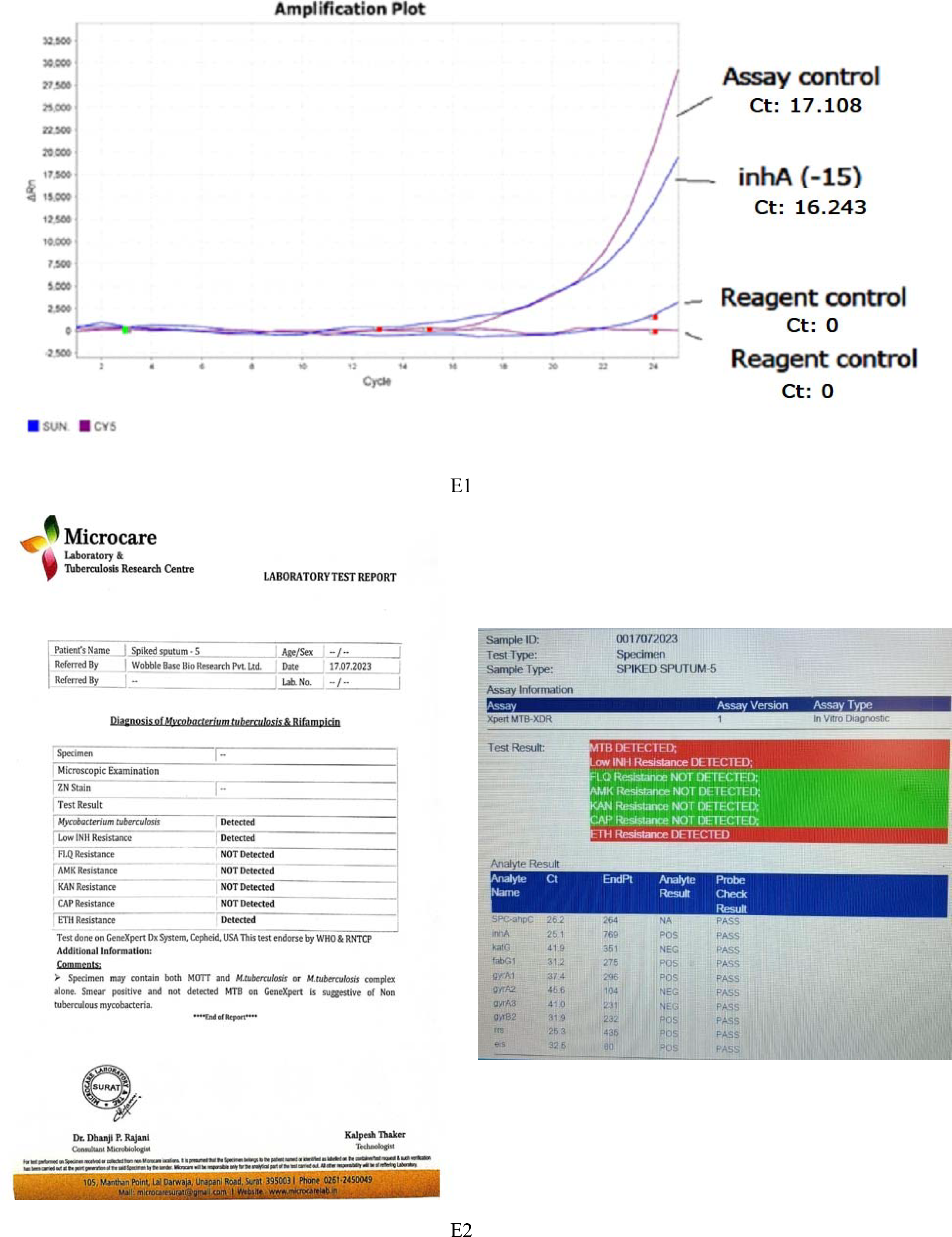
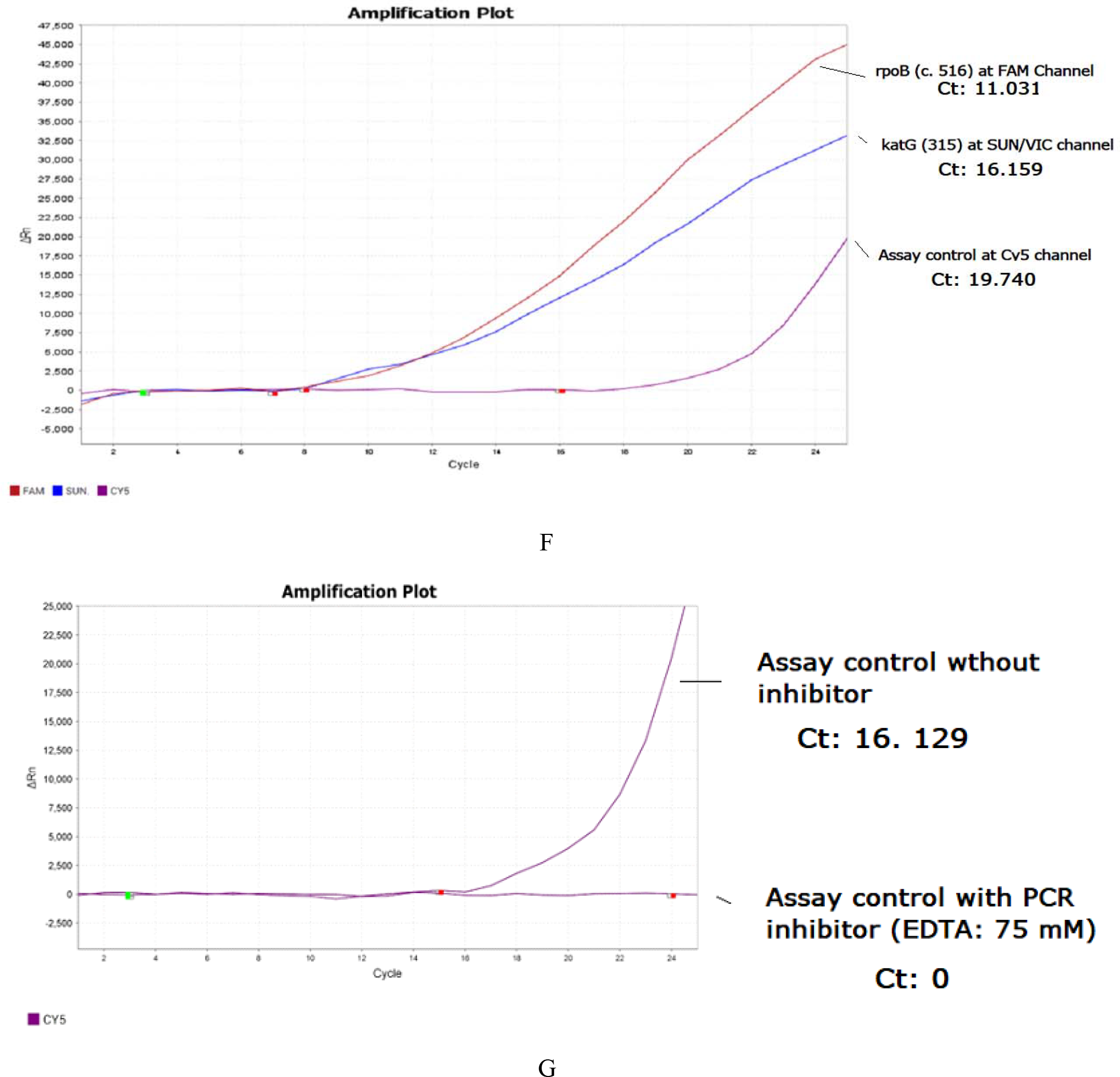
(Panels A1–A2 to E1–E2, F, and G). Real-time PCR assay validation using synthetic mutation templates and integrated internal controls. Panel A1 shows amplification of the *rpoB* codon 516 (GAC→GTC) mutation using the QuantStudio 5 platform, with Panel A2 presenting the parallel GeneXpert MTB/RIF (CB-NAAT) result confirming mutant background at the same locus. Panel B1 shows amplification of the *rpoB* codon 526 (CAC→TAC) mutation, and B2 shows GeneXpert confirmation of the mutant background at codon 526. Panels C1 and C2 respectively demonstrate amplification of the *rpoB* codon 531 (TCG→TTG) mutation and GeneXpert verification of its mutant background. Panel D1 shows amplification of the *katG* codon 315 (AGC→ACC) mutation, with D2 showing its corresponding GeneXpert confirmation. Panel E1 displays amplification of the *inhA* promoter 15 (C→T) mutation, and E2 shows GeneXpert confirmation of mutant background at that site. Panel F demonstrates successful dual-mutation detection of *katG* codon 315 and *rpoB* codon 516 within a multiplexed reaction. Panel G illustrates assay performance under standard conditions and in the presence of a PCR inhibitor (75LJmM EDTA). Panels A1–E1 represent real-time PCR amplification profiles generated on the QuantStudio 5 platform, while A2– E2 correspond to matched CB-NAAT (GeneXpert MTB/RIF) results confirming the intended mutant background of the template DNA. Target-specific details and reaction templates are provided in Table 3.

### Analytical Sensitivity, Amplification Efficiency, and Limit of Detection

The real-time PCR assays developed in this study demonstrated robust performance with consistent amplification of synthetic mutant targets in the presence of wild-type genomic DNA. Mean cycle threshold (Ct) values for mutant target detection ranged from 24.2 to 27.8, depending on the mutation and fluorophore channel used. The internal control (IC) consistently amplified in the Cy5 channel across all reactions with a mean Ct of 28.5 ± 1.2, indicating no inhibitory effects or reagent interference throughout the assay runs.

**Table 3:** Description of DNA template combinations used during validation runs of the multiplex real-time PCR assay incorporating synthetic mutant controls and internal assay controls. Each reaction included wild-type Mycobacterium tuberculosis genomic DNA spiked with mutation-specific synthetic linear DNA fragments at a mean mutant-to-wild-type ratio of approximately 1:70. Panels A1–E1 correspond to reactions targeting single mutations in rpoB (codons 516, 526, 531), katG (codon 315), and the inhA promoter (–15), respectively. Panels A2–E2 show GeneXpert validation of the wild-type status of the genomic DNA background used. Panel F represents a dual-mutation template combining synthetic controls for katG codon 315 and rpoB codon 516. Panel G illustrates assay performance with and without the presence of a PCR inhibitor (EDTA, 75 mM final concentration). Refer to Figure 1 for real-time PCR run images showing amplification of these targets. All runs included a separate, independent reaction without template input, marked in the image as “reagent control,” to monitor assay specificity and reagent integrity.

| Figure No. | Panel No. if any | Description* |
| --- | --- | --- |
| 10 | A1 | *M. tuberculosis* DNA** mixed with synthetic control for *rpoB* (c. 516) mutation + assay control template |
|  | A2 | Sample described in 12-A1 processed by geneXpert test. |
|  | B1 | *M. tuberculosis* DNA** mixed with synthetic control for *rpoB* (c. 526) mutation + assay control template |
|  | B2 | Sample described in 12-B1 processed by geneXpert test. |
|  | C1 | *M. tuberculosis* DNA** mixed with synthetic control for *rpoB* (c. 531) mutation + assay control template |
|  | C2 | Sample described in 12-C1 processed by geneXpert test. |
|  | D1 | *M. tuberculosis* DNA** mixed with synthetic control for *katG* (c. 315) mutation + assay control template |
|  | D2 | Sample described in 12-D1 processed by geneXpert test. |
|  | E1 | *M. tuberculosis* DNA** mixed with synthetic control for *inhA* (-15) mutation + assay control template |
|  | E2 | Sample described in 12-E1 processed by geneXpert test. |
|  | F | *M. tuberculosis* DNA** mixed with synthetic control for *katG* (315) + *rpoB* (c. 516) mutation + assay control template. |
|  | G | Assay control with and without inhibitor (Inhibitor: 75 mM EDTA - final concentration) |

*\*In all cases (Panels A1–E1), the reagent controls were reactions with no template added (No Template Control or NTC) to track possible contamination of the assay reagents, except for Panel F and Panel G. \*\* M. tuberculosis DNA used was confirmed to be of wild type with respect to (1) rpoB codon 516, (2) rpoB codon 526, (3) rpoB codon 531, (4) katG codon 315, and (5) inhA –15 promoter position, respectively. The mean proportion of synthetic mutant to wild-type M. tuberculosis allele used in the runs was approximately 1:70.*

Serial dilutions of synthetic mutant DNA (in a constant background of wild-type MTB DNA) were performed to determine the limit of detection (LOD) for each mutation-specific assay. The LOD was defined as the lowest copy number at which ≥95% of replicates (n = 20) yielded positive amplification with the expected fluorescence signal and no cross-signal in alternate channels.

The observed LOD (limit of detection) values ranged from 25 to 50 copies per reaction, depending on the target. For *rpoB* c.516 (GAC→GTC), the LOD was 25 copies per reaction. The *rpoB* c.526 (CAC→TAC), *katG* c.315 (AGC→ACC), and *inhA* –15 (C→T) mutations each showed an LOD of 50 copies per reaction, while the *rpoB* c.531 (TCG→TTG) mutation had an LOD of 25 copies per reaction.

All assays displayed high amplification efficiency, ranging between 92% and 104%, with R^2^ values >0.995 based on standard curves generated from log-linear dilutions of synthetic templates. No cross-reactivity was observed between wild-type and mutant alleles under optimized blocker-to-primer ratios, confirming the assay’s allele-specific nature and the effectiveness of the wild-type allele blockers.

These results collectively validate the high sensitivity and specificity of the assay platform, with consistent performance even at low mutant-to-wild-type ratios (as low as 1:70), supporting its potential use in early detection of drug-resistant *M. tuberculosis* alleles in clinical settings.

## Discussion

The emergence and spread of multidrug-resistant tuberculosis (MDR-TB) pose one of the most formidable challenges to global TB control programs, especially in high-burden regions (World Health Organization [WHO], 2021). Rapid and precise detection of resistance-conferring mutations in *Mycobacterium tuberculosis* (MTB) is critical for timely intervention and appropriate therapeutic decision-making. In this study, we developed and validated a novel allele-specific real-time PCR platform that employs synthetic mutation controls, 3′-blocked wild-type allele blockers, and contact-quenching probes to achieve high sensitivity and specificity for five clinically significant mutations within the *rpoB, katG*, and *inhA* genes.

A key strength of the approach lies in the successful synthesis and sequence confirmation of DNA controls representing resistance-associated mutations: *rpoB* codons c.516 (GAC→GTC), c.526 (CAC→TAC), and c.531 (TCG→TTG); *katG* codon 315 (AGC→ACC); and *inhA* promoter –15 (C→T). These mutations are among the most prevalent globally and are well-documented markers for resistance to rifampicin and isoniazid, the cornerstone drugs in first-line anti-TB therapy (Zaw et al., 2018; Miotto et al., 2017). The SOEing technique used to generate these controls allowed precise in vitro mutagenesis and recombinant template creation without reliance on restriction enzymes, offering a scalable and cost-effective alternative to plasmid-based standards (Horton et al., 1990).

The functional evaluation of 3′-blocked wild-type allele blocker probes confirmed their ability to suppress amplification of non-mutated templates without affecting mutant allele detection. This strategy significantly improved allele discrimination, an essential requirement in samples with low-frequency mutant populations, common in early or mixed infections (Cohen et al., 2012). Importantly, our real-time PCR results showed no inhibition from the blockers, affirming their compatibility with multiplex qPCR and contact-quenching probe chemistry.

Analytical validation demonstrated that all assays were capable of reliably detecting mutant templates even in the presence of an excess of wild-type DNA (1:70 ratio), simulating conditions often encountered in sputum or extrapulmonary samples. This level of sensitivity aligns well with current international diagnostic benchmarks. The determined limits of detection (LOD), ranging from 25 to 50 copies per reaction, are comparable to those reported for commercial platforms such as GeneXpert MTB/RIF (Boehme et al., 2010), while offering greater flexibility in mutation panel customization.

Mean Ct values (24.2–27.8 for mutant targets and 28.5 ± 1.2 for the internal control) indicated robust amplification kinetics. The high amplification efficiencies (92% to 104%) and strong linearity (R^2^ > 0.995) across standard curves further validate the quantitative reliability of the platform. The assay’s performance was consistent even under multiplexed conditions, as demonstrated by the successful dual detection of *rpoB* and *katG* mutations within a single reaction (Figure 10F), which is advantageous for clinical workflows.

Notably, internal amplification controls (IACs) performed reliably in all reactions and under inhibitor-challenged conditions (EDTA at 75 mM), supporting the assay’s robustness for clinical applications (Figure 10G). This is especially important for resource-limited settings, where sample quality and handling conditions may be variable.

Parallel testing using the GeneXpert platform confirmed that the wild-type *M. tuberculosis* DNA used in these experiments was mutation-free at the relevant loci, strengthening the credibility of the synthetic control-based assay evaluations (Figure 10A2-E2).

Compared to commercial molecular tests like GeneXpert MTB/RIF and Hain’s Line Probe Assays (LPA), the method described here offers two major advantages: (1) customizability, allowing inclusion of emerging or region-specific mutations; and (2) lower assay cost and equipment requirement, potentially enabling broader implementation in decentralized settings (Nikolayevskyy et al., 2016).

In summary, the findings from this study provide compelling evidence for the clinical utility of a contact-quenching, allele-specific qPCR system that leverages synthetic mutation standards and wild-type blocking probes. The platform demonstrated high analytical sensitivity, specificity, and reproducibility, even under multiplexed and inhibitor-rich conditions. Given the growing threat of drug-resistant TB, especially in high-burden regions like India and Sub-Saharan Africa, such technologies could serve as critical components in national and global TB control strategies.

While the results from this study demonstrate the high analytical performance of the allele-specific qPCR platform with contact-quenching detection and synthetic controls, several limitations must be acknowledged.

First, the assay was validated using synthetic DNA templates and wild-type genomic DNA under controlled laboratory conditions. Although this approach provides a rigorous and reproducible foundation for analytical validation, it does not fully capture the complexity and variability of clinical specimens, such as sputum samples, which often contain PCR inhibitors, variable bacterial loads, or degraded DNA. Future studies must evaluate assay performance using a large cohort of de-identified clinical isolates, including smear-positive and smear-negative samples, to assess diagnostic accuracy in real-world settings.

Second, while the selected mutations (*rpoB* codons 516, 526, 531; *katG* 315; *inhA* –15) represent the most prevalent MDR markers globally, they do not encompass the full spectrum of drug resistance-conferring mutations. Resistance to other drugs such as ethambutol, fluoroquinolones, and second-line injectable agents was not addressed in this platform. The assay’s design, however, allows for modular expansion. Incorporating emerging or region-specific resistance markers into the multiplex panel, such as *embB, gyrA*, and *rrs* gene mutations will be critical for future iterations.

Third, while the 3′-blocked allele blocker probes showed high specificity, the possibility of off-target suppression or partial blocking in samples with complex sequence heterogeneity warrants further investigation. Assay optimization in diverse genomic backgrounds, including mixed-strain infections and minority variant populations, should be prioritized.

Lastly, although contact-quenching detection is cost-effective and highly specific, it is not yet as widely adopted or standardized as conventional hydrolysis-based probe chemistries. Broader adoption will depend on demonstrating equivalence or superiority across multiple assay platforms and laboratories.

Future work will focus on expanding the assay to include additional mutations relevant to pre-XDR and XDR-TB, validating the assay on a multicentric clinical sample set, and comparing its performance with WHO-endorsed diagnostic platforms such as GeneXpert MTB/RIF Ultra and Hain’s Line Probe Assay Version 2. In addition, development of a lyophilized, cartridge-based version of the assay for point-of-care application is under consideration, with the goal of integrating it into national TB control programs, particularly in resource-limited settings.

## Conclusion

This study presents the successful development, optimization, and analytical validation of a multiplex real-time PCR assay for the detection of key multidrug-resistant (MDR) mutations in *Mycobacterium tuberculosis*. Using synthetic mutant DNA controls generated via Splicing by Overlap Extension (SOEing), along with 3′-blocked wild-type allele-specific blockers and contact-quenching fluorescence detection, the assay demonstrated high sensitivity, specificity, and robustness. The platform reliably detected five clinically important mutations in *rpoB* (codons 516, 526, 531), *katG* (codon 315), and *inhA* promoter (–15), even in the presence of a large excess of wild-type DNA.

The inclusion of an internal control, compatibility with multiplexed formats, and the ability to detect low-copy mutant alleles (LOD: 25–50 copies per reaction) underscore the assay’s utility in early and accurate resistance detection. Its flexibility and cost-effectiveness, compared with existing commercial assays, further support its potential for implementation in diagnostic laboratories, especially in resource-limited, high-TB burden settings.

Future work will focus on expanding the mutation panel, validating the assay in clinical samples, and adapting the system into a field-deployable format. Overall, this platform offers a promising molecular diagnostic tool for the timely and precise detection of MDR-TB, contributing to more effective TB control strategies globally.

## Data Availability

All data produced in the present study are available upon reasonable request to the authors

## Acknowledgement

The authors gratefully acknowledge Microcare Laboratory and Tuberculosis Research Center, Surat for their invaluable support in providing clinically characterized *Mycobacterium tuberculosis* isolates and genotypically profiled genomic DNA. Special thanks to SN GeneLab Pvt Ltd for assisting in sequencing the synthetic linear DNA fragments generated using SOEing.

We also acknowledge the generous funding support from the Biotechnology Industry Research Assistance Council (BIRAC) under the Small Business Innovation Research Initiative (SBIRI) scheme, which enabled the successful execution of this study.

